# Characterizing the Extended Language Network in Individuals with Multiple Sclerosis

**DOI:** 10.1101/2023.08.30.23294843

**Authors:** Alexander S. Ratzan, Leila Simani, Jordan D. Dworkin, Korhan Buyukturkoglu, Claire S. Riley, Victoria M. Leavitt

## Abstract

**Background:** Language dysfunction is increasingly recognized as a prevalent and early affected cognitive domain in individuals with MS.

**Objectives:** To establish a network-level model of language dysfunction in MS.

**Methods:** Cognitive data and 3T functional and structural brain MRI were acquired from 54 MS patients and 54 matched healthy controls (HCs). Functional summary measures (anteriority, segregation, betweenness, within-ness) of the extended language network (ELN) were calculated and structural imaging metrics were derived. Group differences in ELN connectivity were evaluated. Associations between ELN connectivity and language performance were assessed; in the MS group, an unsupervised learning approach was used to assess relationships between multimodal neuroimaging features derived from language-related areas and performance on language tasks.

**Results:** The MS group performed worse on semantic fluency and rapid automized naming tests (*p*<0.005) compared to HC. Regarding ELN measures, the MS group exhibited higher within-ELN connectivity than HCs (*p*<0.05). Principal component analysis (PCA) yielded a multimodal latent component that uniquely correlated with language performance (*p*<0.05).

**Conclusion:** We identified network-level functional and structural measures to potentially characterize language dysfunction in MS. Further studies leveraging these features may reveal mechanisms and predictors of language dysfunction specific to MS.

## Introduction

Cognitive impairment is common in multiple sclerosis (MS).^1^ While memory and processing speed are considered the most affected domains,^2-4^ language dysfunction is prominent.^5-8^ Recently, a test of rapid automatized naming was the only objective measure (of 9) distinguishing recently diagnosed patients with (pwMS) from matched healthy controls (HCs), and word finding difficulties were the most common cognitive issue of pwMS early in their disease course.^9^ Another recent study found that a measure of language fluency in combination with the Symbol Digit Modalities Test was the most discriminative summary measure (compared to the Brief International Cognitive Assessment in Multiple Sclerosis, BICAMS) and showed decline (rather than improvement) over 1-year follow-up.^10^ The field lacks a mechanistic model of language dysfunction in MS.

Imaging studies investigating language impairment traditionally focus on one-to-one brain-behavior relationships. In MS, the few imaging studies to date evaluating language examined cortical thickness and white matter microstructure.^9,11^ Developing a more comprehensive network-level model of language for MS will permit mechanistic insights into this functionally limiting cognitive deficit. The extended language network (ELN) derived from resting-state functional magnetic resonance imaging (fMRI) of 970 healthy adults, has been shown to be reproducible during resting-state and task-based fMRI, recommending its use as a relevant network for exploring language dysfunction.^12-15^

We utilized the ELN and a previously developed approach for characterizing network reorganization of functionally specific subnetworks using resting state functional connectivity (rsFC). Summary measures to quantify non-random patterns of network-level language network reorganization were calculated: within-ELN connectivity, between-ELN connectivity, segregation index (Seg-I), anteriority index (Ant-I).^16-18^ We tested: (a) whether distinct patterns of functional organization of the ELN are observable in pwMS compared to HCs; (b) whether ELN rsFC is associated with language function within the MS group; and (c) whether multimodal language-related neuroimaging measures are uniquely related to language performance.

## Methods

Study procedures were approved by the Columbia University institutional review board. Written informed consent was obtained from all participants.

### Participants

For the MS group, we utilized data from a prospective cohort of adults diagnosed with relapsing-remitting MS.^18,19^ A separate sample of age, sex, and IQ matched healthy adults served as a comparison group.^20^ For sample characteristics, see Table 1.

### Cognitive measures

All participants completed the following language tests: the Controlled Oral Word Association Test (COWAT): phonemic fluency (FAS), semantic fluency (Animals); rapid automatized naming: Stroop Word Naming, Stroop Color Naming. The MS group completed the Brief Visuospatial Memory Test-Revised (BVMT-R) and the Symbol Digit Modalities Test (SDMT).

### MRI data acquisition

In the MS sample, images were acquired on a 3 Tesla MR scanner (GE Discovery) employing the following parameters: Structural images: T1-weighted BRAVO 1 mm sequence, TE/TR=2.7, 7200 ms, voxel resolution=1×1×1mm^3^. Functional images: echo planar imaging (EPI), 66 axial slices, TE/TR=25, 850 ms, voxel resolution = 2×2×2 mm^3^. During the 9-minute resting-state scan acquisition, participants were instructed to remain still and awake, with eyes closed. In HCs, images were acquired on a 3 Tesla MR scanner (Philips Achieva Magnet). Structural images: T1-weighted magnetization-prepared rapid gradient-echo (MPRAGE) scan, TE/TR=3, 6500 ms, voxel resolution of 1×1×1 mm^3^. Functional images: echo planar imaging, 41 axial slices, TE/TR=20, 2000 ms, voxel resolution = 2×2×2 mm^3^. During the 7- minute resting-state scan acquisition, participants were instructed to remain still and awake, with eyes closed ^21-23^.

### Resting-state functional connectivity (rsFC)

Functional connectivity analysis was performed using the CONN toolbox (version 21a; http://www.nitrc.org/projects/conn), implemented in MATLAB 2021a (MathWorks Inc., Natick, MA, USA). Images were preprocessed using CONN toolbox default preprocessing pipeline. Functional data were spatially realigned, unwarped, and slice-time corrected. Outlier scans were identified using conservative thresholds (framewise displacement above 0.5 mm or global blood oxygen-level-dependent (BOLD) signal changes greater than Z = 5). Functional and anatomical images were then normalized into the common stereotaxic Montreal Neurological Institute (MNI) space with 2-and 1-mm isotropic voxels, respectively, and segmented into gray matter, white matter, and cerebrospinal fluid (CSF) tissue classes. Finally, functional data were spatially smoothed using an 8mm full width half maximum (FWHM) Gaussian kernel. Successful normalization and smoothing of functional and anatomical images were confirmed manually for each subject. Next, functional images were denoised using the CONN toolbox default denoising pipeline. Confounding effects were estimated and removed from the BOLD signal for each voxel for each subject using the default anatomical component-based noise correction procedure (aCompCor). Finally, a bandpass filter (0.008, 0.09 Hz) was applied to functional data to investigate low-frequency BOLD signal fluctuations while minimizing influence of physiological and head-movement noise.

### rsFC processing

Regions of interest (ROIs) were defined based on the default CONN toolbox atlas, the Schaefer 200-parcel parcellation,^24^ and the ELN atlas. The default CONN toolbox atlas includes 132 cortical, subcortical, and cerebellar ROIs from the Harvard-Oxford Atlas and Automated anatomical labelling (AAL) atlas 3.^25^ The 17 network Schaefer 200-parcel parcellation was extracted in the 2mm space.^26^ A custom atlas was created for the ELN by importing spherical ROIs using the MNI coordinates specified for each region by Tomasi & Volkow.^12^ Spherical ROIs were utilized consistent with prior work examining language network perturbations in normative and clinical populations^27-29^, resulting in a 23 ROI atlas of the ELN (Figure 1).

**Fig. 1:**
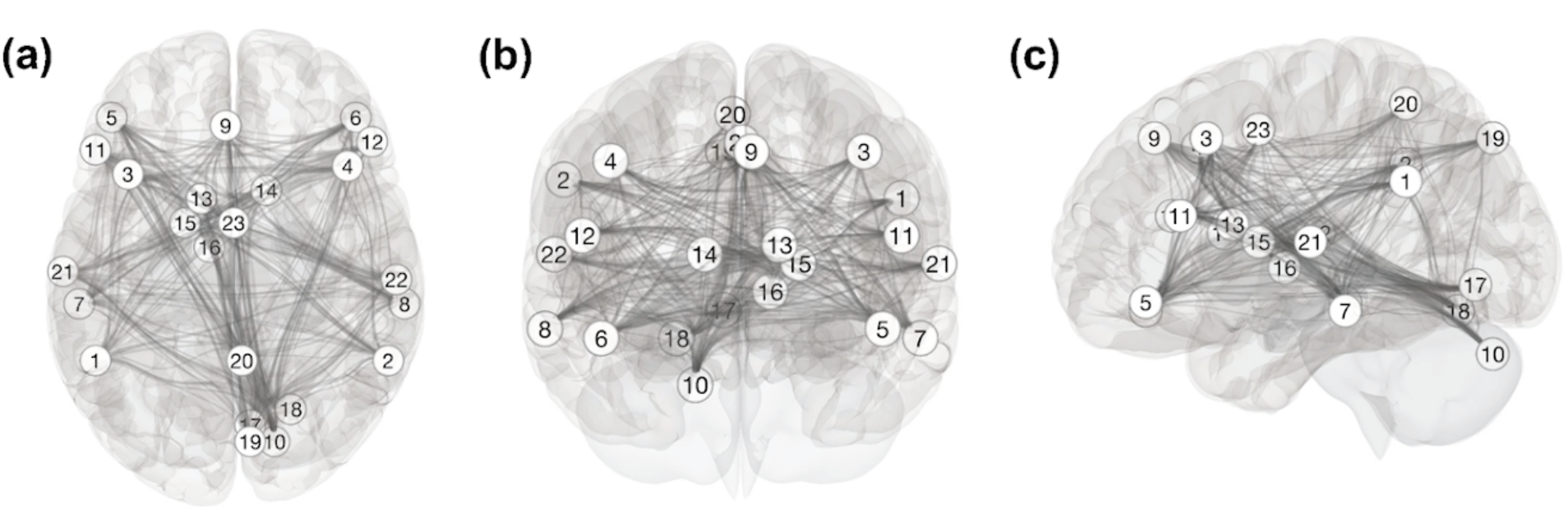
Nodes and connections of the ELN defined by Tomasi & Volkow, 2012: (a) axial view, (b) coronal view, (c) left sagittal view. 1= Wernicke’s area; 2= Right inferior parietal; 3= Middle frontal; 4= Pars opercularis; 5= Left pars orbitalis; 6= Right pars orbitalis; 7= Left inferior temporal; 8= Right inferior temporal; 9= Superior frontal; 10= Cerebellum; 11= Broca’s area; 12= Pars triangularis; 13= Left caudate; 14= Right caudate; 15= Putamen/globus pallidus; 16= Ventral thalamus; 17= Striate; 18= Extrastriate; 19= Posterior parietal; 20= Superior Parietal; 21= Left superior temporal; 22= Right superior temporal; and 23= Cingulate.

For each participant, Pearson’s correlation coefficients were calculated for all possible pairs among ELN and non-ELN regions of the brain (Schaefer 17 network 200-parcel parcellation). A 23×23 connectivity matrix of Fisher z-transformed r-values for each participant was thus derived for the ELN, and a 23×200 connectivity matrix of Fisher z-transformed r-values was derived for the connectivity values between ELN and non-ELN nodes. Prior to calculating the ELN summary measures, the matrices were passed through neuroCombat, a site-harmonization tool to reduce scanner effects introduced by between-group differences. This method estimates an additive and a multiplicative site-effect coefficient at each parcel, thus accounting for regional scanner differences. neuroCombat was successfully applied to mitigate scanner differences in previous fMRI studies allowing for harmonization of data collected across multiple scanners.^30-32^ The diagonal and negative values of the matrices were set to 0 in the final matrices permitting only positive interactions between ROIs to contribute to derived measures of network interactions.

### ELN summary measures

Four ELN connectivity measures were calculated from the resultant matrices: within-ELN (average connectivity of nodes within the ELN); between-ELN (average connectivity between nodes of the ELN and nodes of the rest of the brain); Segregation Index (Seg-I, relationship of within-ELN connectivity to between-ELN connectivity); and Anteriority Index (Ant-I, average connectivity of 11 anterior nodes divided by average connectivity of 12 posterior nodes of the ELN).^17,33^ Higher Seg-I indicates greater ‘within-ness’ than ‘between-ness’, that is, greater reliance on within-ELN connections compared to connections between ELN and the rest of the brain. Ant-I captures differences in connectivity of anterior and posterior regions of the ELN. An Ant-I value of 1 indicates equivalent connectivity of anterior and posterior regions, higher Ant-I indicates increasing reliance on anterior regions, and lower values indicate greater reliance on posterior regions. See Figure 2 for a graphical representation of the ELN measures.

**Fig. 2:**
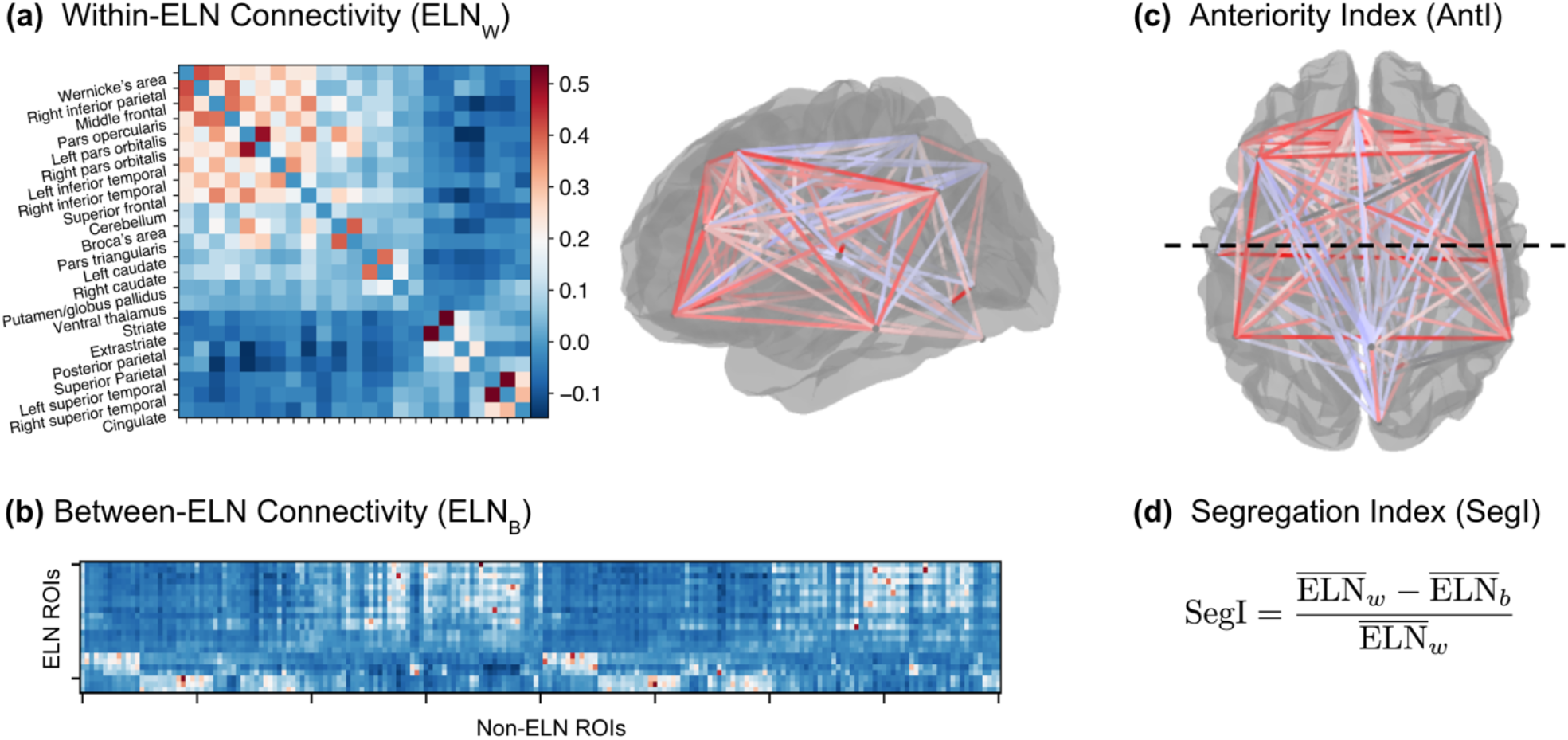
Derivation of ELN summary measures. (a) rsFC matrix of the 23 ELN nodes used to calculate mean within-ELN connectivity. Left sagittal view of within-ELN connectivity. Darker red edges represent stronger connectivity between nodes (higher r-values) while lighter red and light blue represent low connectivity. (b) rsFC matrix of the 23 ELN ROIs with 200 non-ELN ROIs used to calculate mean between-ELN connectivity. Note that negative connectivity values are set to zero in the calculation of ELN summary measures. (c) Connectivity between the anterior and posterior nodes (separated by dotted line) of the ELN. AntI is computed as the mean connectivity among the anterior nodes divided by the mean connectivity among the posterior nodes. (d) Segregation index computed as the difference of mean within-ELN connectivity and mean between-ELN connectivity normalized by mean within-ELN connectivity.

### Structural imaging measures

Additional imaging measures were extracted from structural T1 images of the MS cohort. Cortical thickness of 68 cortical regions (34 per hemisphere) was calculated from manually lesion in-painted 3D-T1 images using Free Surfer (V-6.0) with default settings.^34^ Lesion segmentation was performed using automated Lesion Segmentation Tool (LST).^35^

### Diffusion weighted imaging measures

Raw diffusion-weighted images were corrected for distortions caused by motion, eddy current, and field inhomogeneity using FMRIB’s Diffusion Toolbox within FSL 6.0.4. Probabilistic distribution of 18 major diffusion weighted white matter tracts in each participant were extracted using Free Surfer V-6.0, TRACULA.^36^ Average fractional anisotropy (FA) and mean diffusivity (MD) for each tract were calculated.

## Statistical analyses

One-tailed t-tests assessed group differences in language performance. Two-tailed t-tests assessed whether ELN summary measures were differentially expressed between groups. Pearson’s correlation coefficients were computed within each diagnostic group to evaluate relationships of connectivity measures to language performance. To assess group differences in pairwise ELN connections, two-tailed t-tests were conducted for each node-node connection with the ELN, as well as each node-node connection from the ELN to the rest of the brain (Schaefer 200-parcel parcellation). In a planned exploratory analysis, we assessed relationships of performance on language and non-language cognitive tests to ELN summary measures and traditional functional / structural measures. All p-values were FDR corrected for multiple comparisons. Statistics were conducted with the scipy.stats package in Python.

### Relationship between language-related imaging measures and language performance in MS

Given the limited sample size of our dataset, a correlative unsupervised learning approach was applied to relate multimodal neuroimaging features to language performance. Prior to conducting the analysis, the feature set was determined using a theoretically and empirically driven approach. The feature set included 4 ELN summary measures, 7 rsFC connections deemed specific to the MS group from the pairwise ELN connectivity analysis, 18 cortical thickness measures from putative language regions as well as regions that differed at the rsFC level, and 16 diffusion tensor imaging measures (mean diffusivity and fractional anisotropy) of putative language pathways.^33^ We calculated mean functional connectivity, cortical thickness, fractional anisotropy, and mean diffusivity to include for reference. Sex, age, and T2 lesion volume were included as features in our analysis. This yielded a 54-subject by 52-feature dataset.

First, univariate correlations were calculated between each imaging feature and the available cognitive tests in the MS group. To assess multivariate patterns, PCA was applied to the feature set, retaining the principal components explaining 90% of the variance. Nine subjects were removed from the PCA analysis due to missing features. To focus on language-related imaging measures, global measures and demographic covariates were removed from the PCA analysis yielding a 45-subject by 45-feature dataset (see Figure S2 for identical analysis including omitted measures). Raw subject data was subsequently projected onto the principal components, reducing to 17 features, and correlations to cognitive test scores were computed. Component loadings were visualized for components with relevant correlations to language (Figure 4b). FDR multiple comparisons correction was applied to all statistical tests. Statistical analysis was performed with sci-kit learn and scipy.stats Python packages.

## Results

### Group differences in cognitive test performance

The MS group performed worse than HCs on semantic fluency (*p*<0.005), Stroop Color Naming (*p*< 0.005), and Stroop Word Naming (*p*<0.001) (Table 2).

### Group differences in ELN summary measures

The MS group showed higher within-ELN connectivity (*p*<0.05) and marginally higher Seg-I compared to HCs, although this did not reach the level of statistical significance (*p*=0.08). No group differences were found for between-ELN connectivity or Ant-I (Table 3).

### Relationship of ELN summary measures to language function

There were no significant relationships between ELN summary measures and language function in either group. However, in the MS group there were several relationships trending towards significance on Stroop Word Naming. Both within- and between-ELN connectivity showed trend-level positive correlations with performance on this test (*r*=0.22, *p*=0.118; *r*=0.24, *p*=0.086, respectively).

### Group differences in pairwise functional connectivity

For exploratory purposes, we evaluated group differences in rsFC among all pairwise ELN connections. Five pairwise connections differed for connections *within* ELN (Figure 3), and two pairwise connections differed for connections *between* nodes of ELN and the rest of the brain. While the above connections did not withstand FDR-correction for multiple comparisons, they were included in further exploratory analysis due to their potential relevance as MS-specific language connections.

**Fig. 3:**
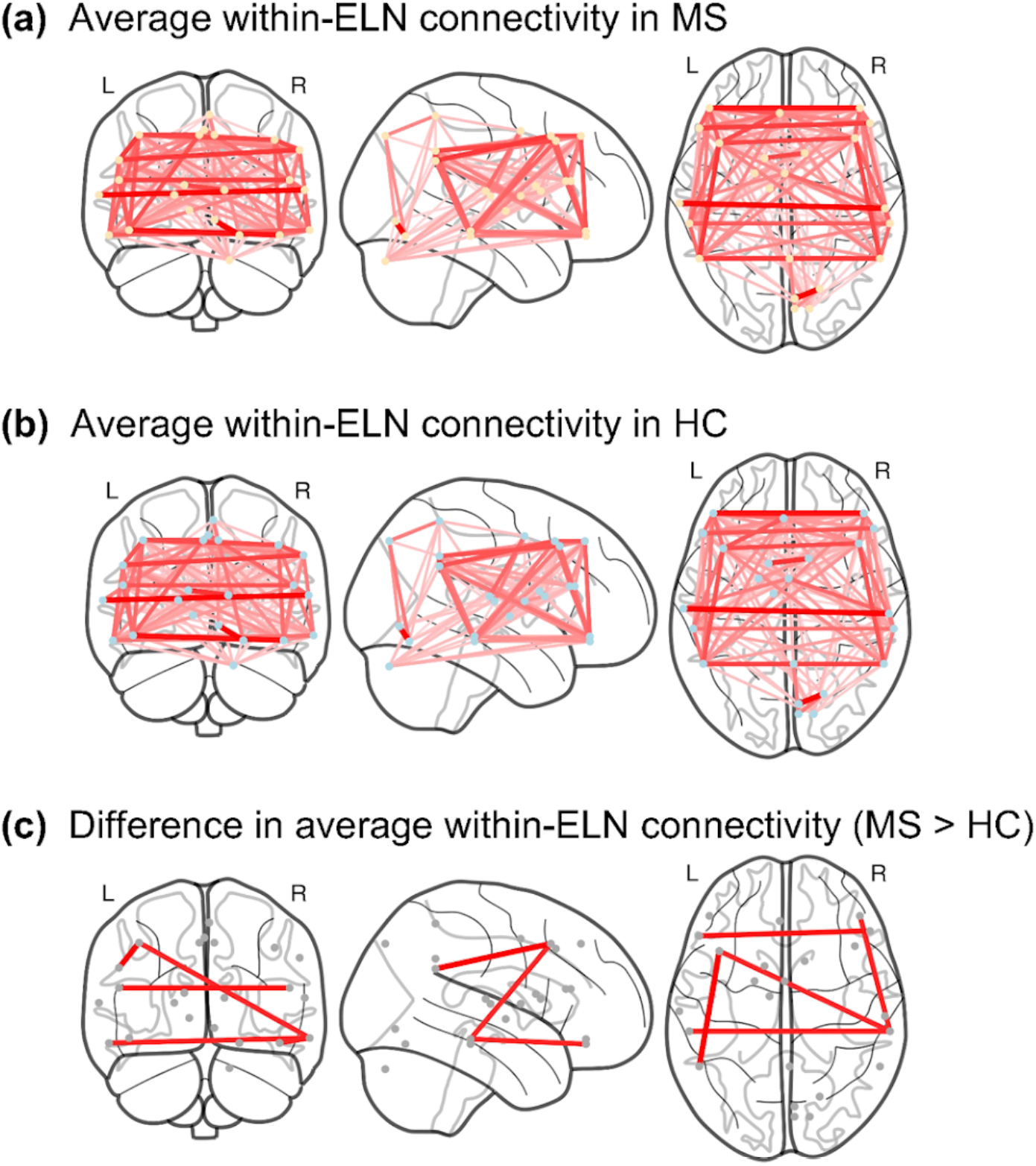
Pairwise ELN rsFC differences between people with MS and healthy controls. (a) pwMS average ELN rsFC. (b) HC average ELN rsFC (c) Difference between pwMS average ELN rsFC and HC average ELN rsFC. Connections include middle frontal to right pars orbitalis, left pars orbitalis to pars triangularis, right pars orbitalis to right inferior temporal, left inferior temporal to right inferior temporal, and broca’s area to pars triangularis.

### Relationship of multimodal language-related imaging measures to language function

Pearson correlations were computed between all functional and structural imaging measures and the language and non-language cognitive tests. Several features correlated with Stroop Words, BVMT, and SDMT, however none withstood multiple comparisons correction (Figure S1). PCA reduced the dataset to 17 latent components, composed of linear combinations of the original features. Correlating the reduced data to cognitive scores unveiled a latent component that significantly correlated with the Animals (semantic fluency) language test (*r*=0.51; *p-uncorrected*<0.0005; *p-corrected*<0.05). None of the latent components significantly correlated with non-language cognition. The top tertile of loadings for this latent component (PC16) are visualized in Figure 4. ELN summary measures, pairwise rsFC, cortical thickness, and diffusion measures are relatively evenly represented in the loadings. Loadings for PC12 are visualized given the potential language specific nature of this component. Ant-I is represented in the top tertile of loadings for PC16 and Seg-I, ELN within-ness, and ELN between-ness are represented in the top loadings of PC12.

**Fig. 4:**
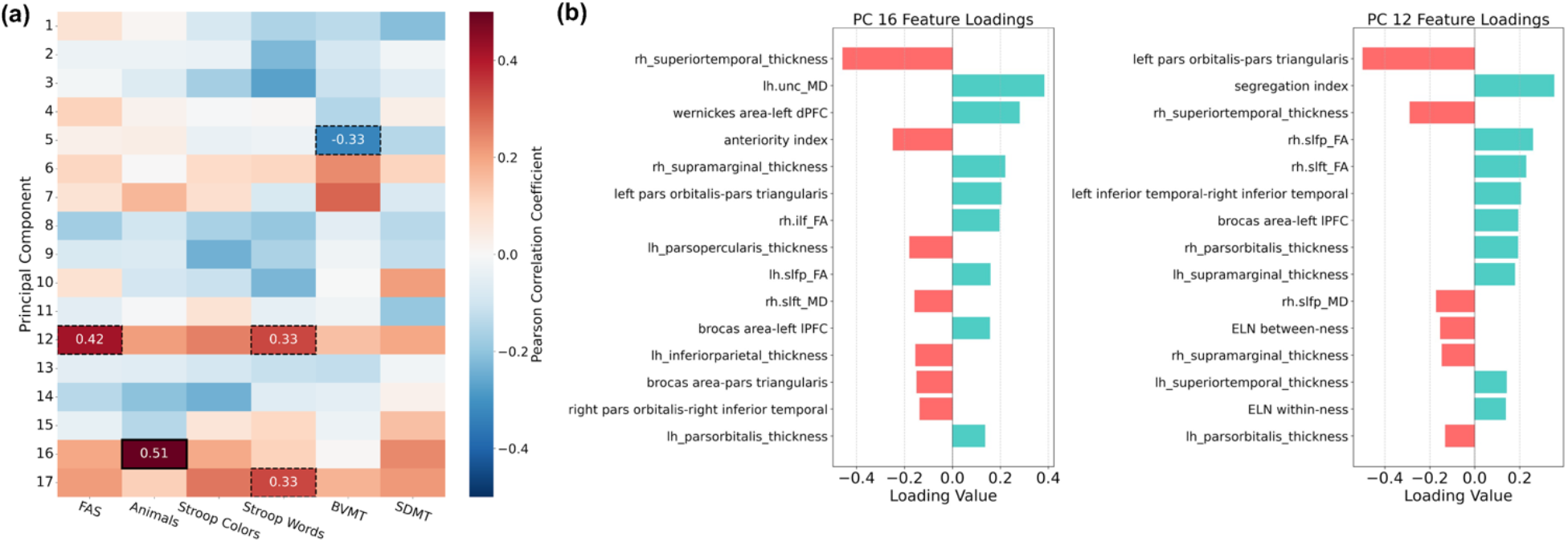
(a) Heatmap showing the correlation between PCA reduced data and performance on four language and two non-language tasks. 45 language-related imaging features are used for PCA. (b) Top tertile of feature loadings for PC16 (*p-corrected<0*.*05*) and PC12 (*p-uncorrected<0*.*05*). Dashed borders denote significance for *p-uncorrected<0*.*05* and solid outlines denote significance for *p-corrected<0*.*05*.

## Discussion

These results reveal decrements in language function and ELN alterations in pwMS. In our approach, we derived summary measures to capture large-scale organizational shifts of the ELN. This approach has been used in prior work to test functionally meaningful shifts in memory subnetwork organization.^17,18^ We found that pwMS exhibited greater within-ELN connectivity relative to HCs. A trend-level difference was observed for Seg-I. While there were no significant associations of index scores to language performance, trend-level associations suggest a lack of statistical power. Finally, our exploratory analysis testing a multimodal model of language function highlighted a latent component composed of diverse neuroimaging features that may uniquely explain language impairment in MS.

Studies of rsFC in MS generally evaluate connectivity within primary brain networks, e.g. default-mode, salience network.^37,38^ Summary measures permit explicit tests of large-scale network reorganization relevant to prespecified cognitive domains. We calculated network-level summary measures of the ELN based on our prior work.^16,18^ Within-ELN connectivity characterizes the intrinsic wiring of the language network, with higher values suggesting stronger ‘local’ processing. Between-ELN connectivity, conversely, captures the affinity for nodes of the ELN to functionally wire with non-ELN cortical regions, a proxy for ‘global’ connectivity. Seg-I describes the balance between ‘local’ and ‘global’ ELN connectivity (Figure 2). Comparing these measures between groups and relating them to behavioral performance provides insights into disease-specific reorganization related to language dysfunction. Our results highlight higher within-ELN connectivity in MS versus HCs, suggesting stronger connectivity of nodes within the ELN. We observed slightly elevated (though non-significant) segregation in the MS group suggesting more local than global connectivity of the ELN. There is moderate agreement of these results with prior work reporting network segregation within functional subnetworks in cognitively impaired pwMS.^37-39^ Segregated processing is hypothesized to represent functional rerouting as a compensatory mechanism to preserve communication between distant and potentially disconnected brain regions.^40^ Some studies have found different patterns of network segregation in MS, potentially due to high sensitivity of the measure to disease stage.^41^ Nonetheless, these studies implicate functional rerouting as a probable phenomenon with relevance for cognitive status throughout the MS disease course and suggest that as neurobiological damage increases, functional reorganization takes place.^42^

Our exploratory analysis sought to address the multimodal nature of language impairment in MS. In lieu of analyzing all candidate brain regions and pairwise connections, we first subset our features to putative language regions and pathways enabling more targeted and theoretically driven hypothesis tests of language function. Univariate correlations showed moderate associations of cortical thickness with language and non-language cognition (prior to multiple comparisons correction), suggesting their involvement in multiple cognitive domains beyond language.

Univariate relationships, however, are limited in capturing the complexity of disease related cognitive impairment. Using PCA, we derived 17 principal components explaining 90% of the variance in the dataset. Each latent component represents an orthogonal basis vector constructed as a linear combination of the original features. By correlating our dimensionality-reduced data to each cognitive test, we identified a latent component (PC16) that was significantly and uniquely correlated with semantic fluency. Observing the loadings of this principal component (PC16), there appears to be an even contribution of network-level, functional, and structural measures, further pointing towards the utility of multimodal neuroimaging biomarkers for characterizing language function. PC12, although non-significant, displayed preferential associations with language tasks while bearing no relationships with non-language cognition, warranting further investigation of its loadings.

With respect to the loadings, all four ELN summary measures were represented in the combined top tertile of features of PC12 and PC16 along with various cortical thickness, pairwise connections, and diffusion measures of language-related pathways, such as the superior longitudinal fasciculus. This diversity is consistent with the notion that brain network rerouting likely occurs structurally and functionally, at varying levels (i.e., node-node and network-level; Figure 4, Figure S1).^36,41^ It must be noted that our unsupervised approach is purely correlative, not predictive, and applying more complex predictive models to larger datasets would more effectively address whether ELN summary measures better explain language function as compared to regional or pairwise measures.

An advantage of summary measures over node-node connections is that they minimize individual inhomogeneities (e.g., global rsFC, lateralization differences) that hinder standardization of neuroimaging metrics for use as clinical trial outcomes.^14^ Seg-I, for example, is more resistant to scanner effects as it compares relative differences in network activations on a within-subject basis. If global rsFC was higher overall in one group, Seg-I would be unaffected as the value is dependent on the relative ratio of within-to between-ELN connectivity. Within and between-ELN connectivity minimize other critical inhomogeneities such as lateralization differences across individuals. These measures, calculated downstream of our neuroCombat harmonization, rely on the connectivity of many nodes in a network reducing the potential effects of individual node-node outliers. Finally, network summary measures are replicable, simple to calculate, and theoretically driven, making them biologically relevant descriptors of cognition.

Our study has limitations. The MS sample was early in disease progression warranting future studies of later-stage patients. Additionally, longitudinal studies relating changes in imaging measures to cognition are warranted. The MS group was a sample of convenience, adopted from a dedicated study of memory mechanisms; sample size may have limited our ability to detect significant relationships, specifically in the regression analysis. Lastly, we applied neuroCombat, which estimates voxel or parcel-level adjustments to eliminate effects directly associated with different scanner types and protocols. Although the method has been validated for use on a limited number of different scanners, utilizing larger cohorts collected across more scanners would yield more effective adjustments based on scanner effects.^37^

Our findings highlight the value of multimodal network-level imaging metrics for understanding language dysfunction in MS. Future studies of the ELN are critical for advancing mechanistic insights and targeted interventions.

## Acknowledgment

The authors thank Lauren Heuer for assistance in preparing the manuscript for publication, and thank the individuals who participated in the study. The authors also thank Yaakov Stern for his generous sharing of data collected for the Reference Ability Neural Networks (RANN) cohort used in this study.

## Data Availability

Data used in this study are available upon request to the corresponding author.

---

**Fig. S1:**
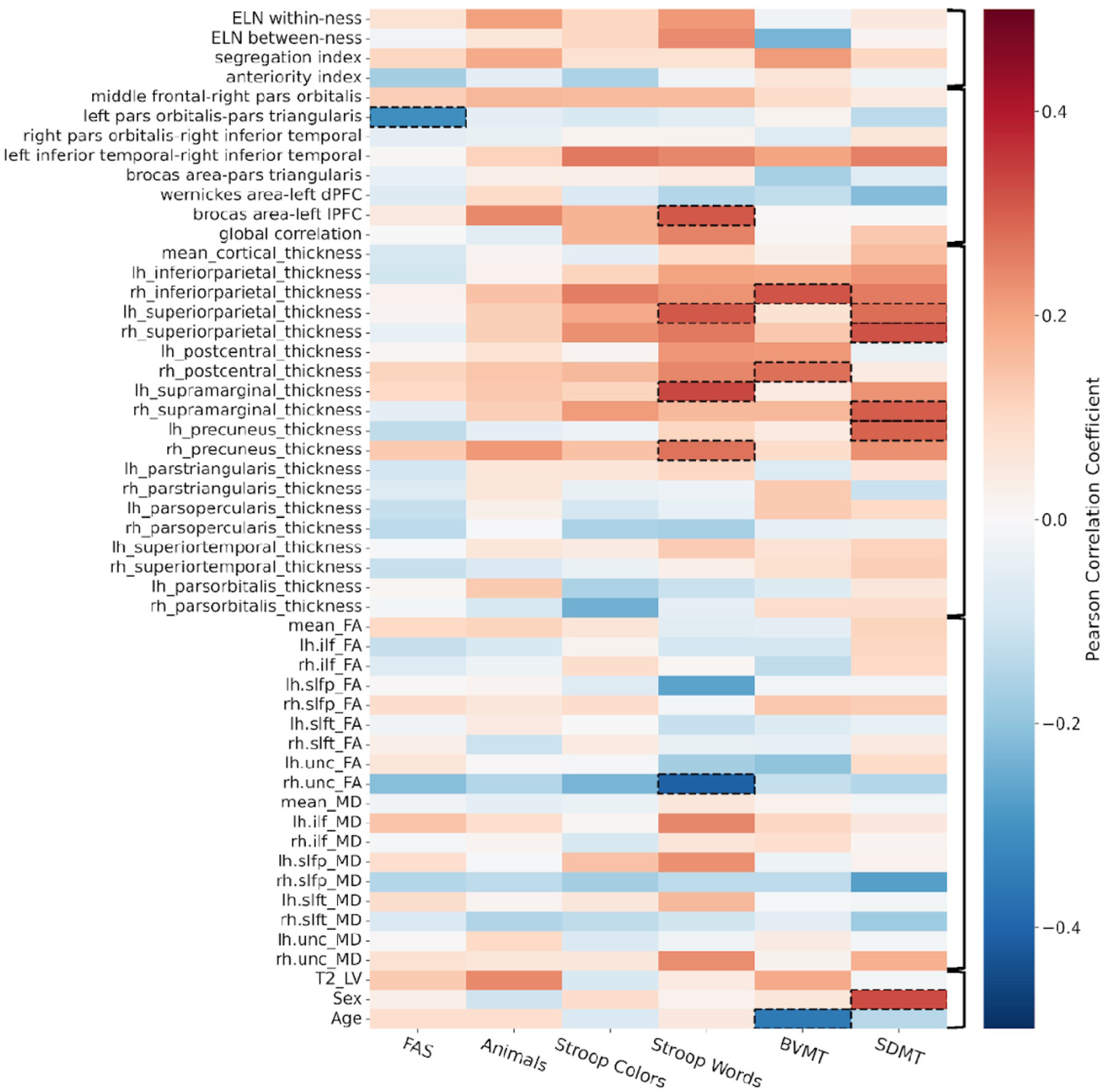
Heatmap for all features correlated with behavioral performance on four language tasks and two non-language tasks. Features are grouped from top to bottom by ELN rsFC summary measures, additional derived rsFC measures, cortical thickness and diffusion measures of putative language areas, and covariates (T2 Lesion Volume, Sex, Age). Dashed outlines indicate p<0.05 for respective correlation coefficients. No relationships withstood FDR correction for multiple-comparisons.

**Fig. S2:**
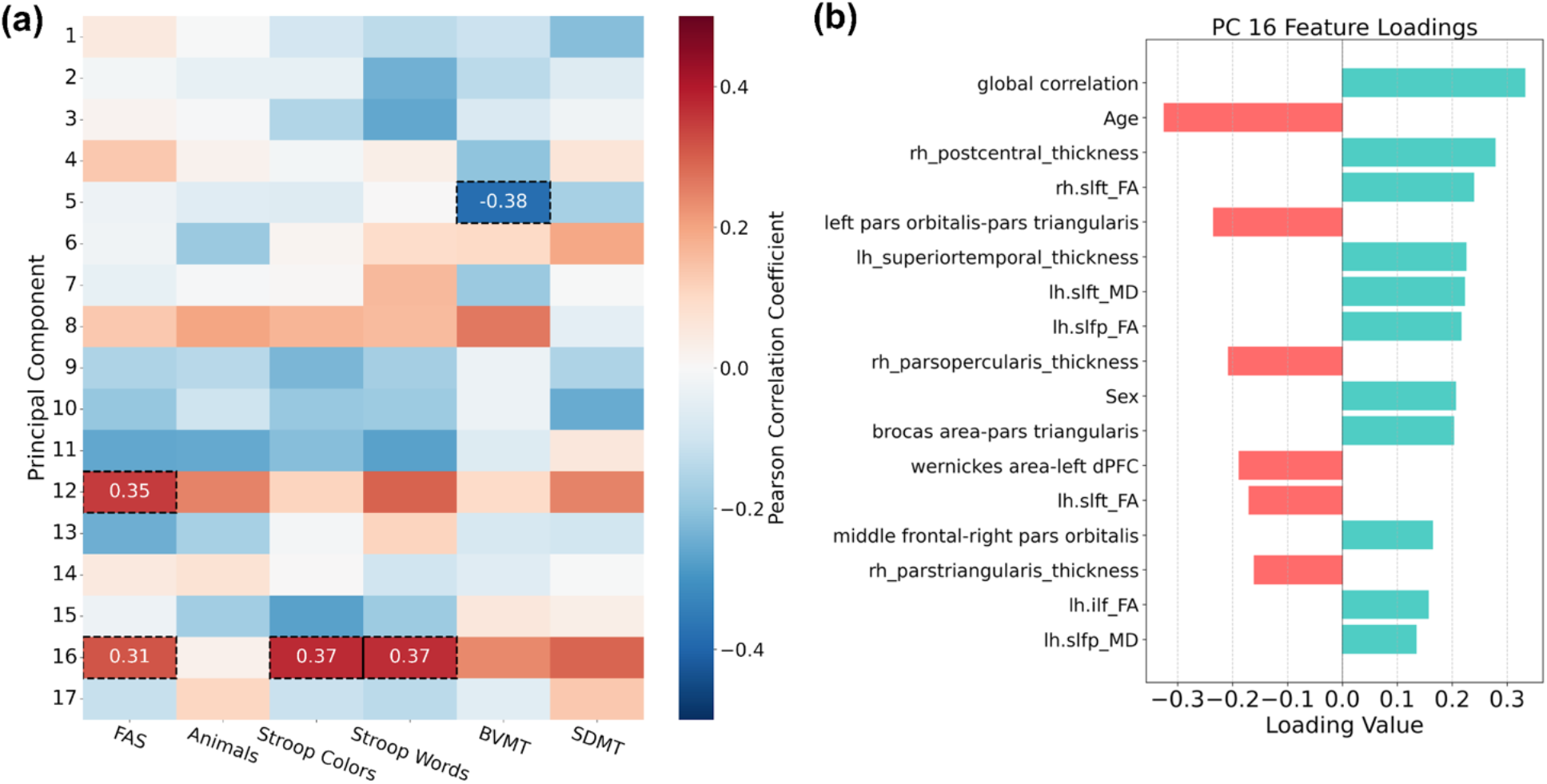
(a) Heatmap showing the correlation between PCA reduced data and performance on four language and two non-language tasks. 45 language-related imaging features are used for PCA along with 5 additional global brain measures and 2 demographic covariates. (b) Top tertile of feature loadings for PC16. Dashed borders denote significance for *p-uncorrected<0*.*05* and solid outlines denote significance for *p-corrected<0*.*05*.

